# Ranking-optimized survival models can underperform fixed-horizon clinical prediction

**DOI:** 10.64898/2026.06.13.26355565

**Authors:** Truong Quynh Hoa, Hoang Dinh Cuong, Luu Duc Trung

**Author notes:** **Corresponding author.** (Truong Quynh Hoa).

## Abstract

Machine-learning survival models are increasingly proposed for intensive-care mortality prediction and are usually judged by the concordance index, a ranking metric averaged over follow-up. Yet many bedside decisions require a probability at a specific time, such as 60- or 180-day mortality. We asked whether ranking-optimized models perform competitively at fixed clinical horizons when compared with attending-physician judgment and the original 1995 SUPPORT logistic model. Reanalyzing the SUPPORT2 cohort (9,105 critically ill adults; five United States centers; 1989-1994) with a stratified 70/15/15 split, we compared a gradient-boosted survival model, the physician’s recorded prognostic estimate, and the 1995 model at 60 and 180 days, and tested several alternative learners. The survival model achieved a competitive ranking concordance (0.705) but underperformed both comparators at fixed horizons: at 60 days its area under the ROC curve was 0.750, versus 0.808 for physicians (on the matched sample) and 0.827 for the 1995 model, a gap reproduced across eight independent splits and statistically reliable after multiplicity correction. Discrimination was equitable across sex, race, and age. Post-hoc recalibration did not change discrimination, so the deficit is not miscalibration. Replacing the ranking objective with timepoint-matched binary training recovered roughly half the gap; neural networks, a deep ranking model, and two timepoint-aware discrete-time models did not close it, indicating an objective-horizon mismatch rather than limited model capacity. Leave-one-disease-out validation revealed severe generalization failure in disease groups absent from training. The physician advantage was conditional on a physician electing to give an estimate; many gave uninformative or no estimate. We recommend reporting timepoint-specific discrimination alongside the concordance index, timepoint-matched training when fixed-horizon predictions drive care, leave-one-subgroup validation, and distribution-free prediction intervals to support selective deployment.

## 1. Introduction

The Study to Understand Prognoses and Preferences for Outcomes and Risks of Treatments (SUPPORT) enrolled 9,105 seriously ill adults at five U.S. medical centers between 1989 and 1994 and produced one of the most widely cited prognostic models in critical care medicine [1]. Three decades later, SUPPORT2 remains a benchmark dataset for survival machine learning, with dozens of papers proposing successively more sophisticated models — Cox elastic net [2], random survival forests [3], gradient-boosted survival ensembles [4,5], and deep learning [6,7,8,9] — all evaluated and compared via the concordance index (C-index) [10].

C-index averages discriminative ability across the entire follow-up distribution. It is appropriate when the clinical question is *how to rank patients by risk*. But many bedside decisions — Should we transfer to the ICU? Should we discuss goals of care? Should we initiate aggressive intervention? — depend on probability estimates at *specific* timepoints, typically a few weeks to several months. A model that achieves an excellent C-index by ranking 1-year survivors above 5-year survivors may not perform well at predicting 60-day mortality, because the inductive bias of its loss function privileges long-horizon distinctions over short-horizon resolution. This phenomenon is well-known in principle [7,8], but its empirical magnitude on benchmark clinical datasets — and particularly in comparison to attending physician judgment recorded at admission — has rarely been quantified rigorously.

SUPPORT2 is uniquely well-suited for such a comparison because it contains physician prognostic estimates (`prg2m`, `prg6m`) collected prospectively at admission. Such measurements are rare in modern electronic-health-record datasets, where physician gestalt is seldom documented numerically. This allows a three-way comparison rarely possible elsewhere: ranking-trained survival ML, the original 1995 logistic regression (a historical reference model), and unaided physician judgment.

In this paper we use SUPPORT2 to answer five linked questions:

1. **Is ranking performance a faithful proxy for timepoint performance?** Comparing GBM Survival’s C-index with its 60- and 180-day AUC against physician estimates and the Knaus 1995 logistic regression.
2. **2. If not, can it be fixed?** Testing post-hoc logistic recalibration (which is rank-preserving and therefore cannot improve AUC) and timepoint-matched binary classification (which can).
3. **3. Does ML generalize across patient subgroups?** Leave-one-disease-out validation across eight disease groups.
4. **4. Can we quantify predictive uncertainty rigorously?** Distribution-free split conformal intervals with empirical coverage validation, including a risk-stratified Mondrian variant.
5. **5. Is the physician advantage robust to missing-data handling?** Sensitivity to missing physician estimates (18.1% cohort-wide; 19.0% on the test set).

The contributions of this paper are: (i) empirical demonstration that ranking-trained survival models (gradient-boosted survival, Cox proportional hazards and elastic net, random survival forest, a deep neural Cox model (DeepSurv), and discrete-time deep survival models (nnet-survival, DeepHit)) underperform timepoint-trained 1995 logistic regression and unaided physician judgment at fixed clinical horizons, with detailed diagnosis of the objective-function mismatch; (ii) a cross-horizon AUC matrix revealing asymmetric generalization; (iii) leave-one-disease-out validation demonstrating severe, statistically reliable failure (BSS confidence interval below zero) in two oncologic subgroups absent from training; (iv) split conformal intervals (marginal + Mondrian) providing distribution-free coverage and supporting selective prediction; (v) sensitivity analysis of the physician comparison to four missing-data imputation strategies, revealing that the apparent physician advantage is partly selection-driven; and (vi) a demographic-fairness analysis (sex, race, age) showing the fixed-horizon deficit is equitable across demographic strata rather than a bias artifact.

### 1.1 Related work

#### Machine learning versus clinicians and legacy scores

A large literature reports that machine-learning *classifiers* match or exceed clinical risk scores and clinician judgment for short-term mortality in emergency and critical care. van Doorn et al. [11], for instance, found gradient-boosted classifiers outperformed internal-medicine physicians and the SOFA/MEDS/REMS scores for 31-day sepsis mortality (AUC 0.85 vs 0.74), and systematic reviews reach broadly similar conclusions. Crucially, this evidence almost always concerns *classification* models trained for a *specific* outcome at a *fixed* time. The contrasting evidence on the SUPPORT cohort itself is older but pointed: the original SUPPORT prognostic model [1,12,13] achieved discrimination roughly equal to physicians’ 2- and 6-month survival estimates (ROC ≈ 0.78), with the best performance obtained by *combining* the two. We revisit that three-way comparison on the modern SUPPORT2 release with ranking-trained survival ML in place of the 1995 logistic, isolating a question the earlier literature did not: does the *training objective* of contemporary survival models change the conclusion at fixed clinical horizons?

#### Limits of the concordance index

The concordance index — the dominant discrimination metric in survival ML — is a pure ranking statistic: invariant to monotone transformations of risk, uninformative about calibration or absolute risk, and dominated by comparisons between patients with near-identical survival times [10,14,15]. Hartman et al. [15] show that a high C-index is difficult to achieve even for genuinely useful time-to-event models, and that the reviewer convention of demanding C-index > 0.7 is poorly grounded. Calibration studies make the converse point: models with strong concordance can emit poorly calibrated probabilities that still reach clinicians [16]. We add a third, deployment-facing failure mode on a real cohort — a competitive C-index (0.705) coexisting with fixed-horizon AUC below both physicians and a 1995 logistic, and with *negative* Brier skill in specific disease subgroups.

#### Objective–horizon mismatch and discrete-time modeling

The mechanism we implicate — optimizing a ranking (Cox partial-likelihood) objective when the deployed task is a probability at a fixed horizon — has a methodological literature. Discrete-time survival models recast prediction as binary classification within time intervals, letting any classifier estimate horizon-specific hazards; Suresh et al. [17] show these can match or exceed the continuous-time Cox model for horizon-specific prediction, and Gensheimer & Narasimhan [8] introduced a scalable neural discrete-time model expressly because Cox-style models must be re-trained to predict at different time points. DeepHit [7] combines ranking and likelihood losses for the same reason. These works argue, on methodological grounds, that the objective matters. We supply the complementary empirical artifact: a controlled, like-for-like demonstration on a clinical cohort that replacing the Cox objective with timepoint-matched cross-entropy recovers roughly half of an otherwise large gap, that the deficit is reproduced by a deep Cox model (DeepSurv) yet not by a timepoint-trained neural network, and that it therefore tracks the objective rather than the learner family.

#### Survival deep learning

Neural survival models — DeepSurv [6], DeepHit [7], nnet-survival [8] — are typically benchmarked by C-index against Cox and random survival forests [3]. We evaluate deep Cox (DeepSurv), discrete-time hazard (nnet-survival), and likelihood-plus-ranking (DeepHit) survival models under fixed-horizon AUC, calibration, and net benefit, and find that neither added capacity nor a likelihood objective rescues fixed-horizon performance — only horizon-matched training does.

#### Distribution-free uncertainty

Conformal prediction yields finite-sample, distribution-free coverage around any model’s outputs [18,19,20,21,22,23,24] and has been extended to right-censored survival data [25]. Risk-stratified (Mondrian) conformal [26] targets coverage valid *within* subpopulations rather than only marginally; clinical applications to date have largely used the marginal variant [27,28]. We use Mondrian conformal to support selective prediction, abstaining where intervals are uninformative.

#### Internal-external and subgroup validation

Leave-one-group-out designs are a recommended form of internal-external validation for transportability [29]. Survival ML benchmarks rarely report subgroup performance with uncertainty; we report leave-one-disease-out Brier skill with confidence intervals and find reliable, below-baseline failure in oncologic subgroups absent from training — failures invisible in the pooled C-index.

#### Contribution and its level

This is not a new algorithm; it is a rigorous empirical reanalysis whose contribution is the *assembly and quantification* of an effect that the methodological literature predicts but had not demonstrated on a clinical cohort with human and historical comparators. We (i) show on SUPPORT2 that ranking-trained survival models — boosted, linear, forest, and deep — underperform attending-physician estimates and a three-decade-old logistic model at 60- and 180-day horizons despite competitive concordance; (ii) isolate the objective–horizon mismatch as a substantial cause via a controlled cross-entropy fix and DeepSurv/MLP dissection; (iii) surface reliable disease-subgroup failures using Brier skill with confidence intervals; (iv) make the physician comparison estimand-explicit; and (v) add risk-stratified conformal prediction and decision-curve analysis for deployment realism. To our knowledge this combination has not previously been assembled on this benchmark. The novelty is therefore one of clinical-methodological synthesis and rigor rather than algorithmic invention — fitting the central, cautionary claim: concordance is not a sufficient basis for fixed-horizon clinical deployment.

## 2. Methods

### 2.1 Data and outcomes

We used the publicly available SUPPORT2 dataset (n = 9,105 patients, 47 columns; https://hbiostat.org/data). The primary outcome was all-cause mortality at fixed clinical horizons of 60 and 180 days, derived from and `death`. At these horizons no patient was censored prior to the horizon (median follow-up 233 days), so binary mortality outcomes were well-defined for the full cohort. For extended-horizon analyses (t = 18, 108, 365 days), we used inverse-probability-of-censoring-weighted (IPCW) AUC [30] as implemented in `sksurv.metrics.cumulative_dynamic_auc`.

### 2.2 Splits and feature processing

Patients were split into training (n = 6,373; 70%), validation (n = 1,366; 15%), and held-out test (n = 1,366; 15%) sets, stratified by mortality status (seed = 42). After cleaning, 43 features remained; pre-computed prognostic outputs (`prg2m`, `prg6m`, `surv2m`, `surv6m`, `aps`, `sps`) were excluded from the feature set and retained as comparator baselines on the test set. The post-discharge variable was excluded as a known leakage variable [31]. Missing values were median-imputed within the training fold; numeric features were z-score normalized using training-fold statistics.

### 2.3 Models

We trained four survival models on the training split: Cox proportional hazards (Cox PH, α = 0.01), Cox elastic net (Cox EN, ℓ1 = 0.5) [2], random survival forest (RSF) [3], and gradient-boosted survival analysis (GBM Survival, n_estimators = 300, learning rate = 0.05, max_depth = 3) [4]. RSF hyperparameters were tuned via grid search on the validation set (max_depth ∈ {None, 10, 15, 20}; min_samples_leaf ∈ {3, 5, 10}; n_estimators ∈ {300, 500}); the best configuration was retrained on combined train+validation.

For the timepoint-matched analysis we trained binary GradientBoostingClassifier models [32] with cross-entropy loss on outcomes at t ∈ {18, 60, 108, 180, 365} days, using the same 43 features and hyperparameters as the survival GBM. As a neural comparator, we additionally trained a multilayer-perceptron classifier (MLP; two hidden layers of 64 and 32 units, L2 = 1e-3, early stopping, seed 42) on the same timepoint outcomes. To probe whether the deficit is specific to tree ensembles, we also trained DeepSurv [6] — a multilayer perceptron (two hidden layers of 64 and 32 units, batch normalisation, dropout 0.2) whose scalar output is a relative-risk score optimised by the Cox partial likelihood, i.e. a deep model sharing the ranking objective. To probe the objective–horizon mechanism directly we also trained two discrete-time deep survival models: nnet-survival [8], which models per-interval conditional hazards with a binary likelihood, and DeepHit [7], which predicts a discrete probability mass function over time bins with a combined likelihood-plus-ranking loss; interval edges included 60 and 180 days so horizon probabilities are exact. All learners (survival GBM, the binary classifiers, the MLP, and DeepSurv) were fit on the training fold only; the validation fold was reserved for conformal calibration, recalibration, and hyperparameter selection, so no model received a training-data advantage in the head-to-head comparisons. We additionally report a five-configuration hyperparameter sensitivity sweep for GBM Survival. Table 1 summarises the complete model and comparator inventory.

**Table 1.** Model and comparator inventory. All learners use the same 43 admission-period features and are trained on the training fold (n = 6,373); the validation fold (n = 1,366) is reserved for conformal calibration, recalibration, and hyperparameter selection; the test fold (n = 1,366) is used only for evaluation. SUPPORT-1995 was developed on the full historical SUPPORT cohort, hence the possible test-set overlap noted in Section 4.4.

| Predictor | Objective / loss | Target | Trained on | Tuning | Role |
| --- | --- | --- | --- | --- | --- |
| Cox PH | Cox partial likelihood | time-to-event | train | fixed | survival baseline |
| Cox elastic net | penalised Cox PL | time-to-event | train | fixed | survival baseline |
| Random survival forest | log-rank splitting | time-to-event | train | grid on validation | survival baseline |
| **GBM Survival** | Cox PL (boosting) | time-to-event (ranking) | train | sensitivity sweep | primary ML model |
| DeepSurv | Cox PL (neural) | time-to-event (ranking) | train | fixed architecture | deep ranking model |
| nnet-survival | discrete-time hazard likelihood | per-interval hazard | train | fixed architecture | timepoint-aware survival |
| DeepHit | discrete likelihood + ranking | time-bin PMF | train | fixed architecture | timepoint-aware survival |
| Binary GBM (per t) | binary cross-entropy | died-by-t | train | as GBM Survival | timepoint fix |
| MLP | binary cross-entropy (neural) | died-by-t | train | fixed architecture | neural timepoint |
| Physician | clinical gestalt | — | — (external) | — | leakage-free comparator |
| SUPPORT-1995 | logistic regression (1995) | 2-/6-mo survival | full SUPPORT cohort | — | historical reference (possible overlap) |

### 2.4 Comparators

The attending physician’s prognostic estimate was extracted as 1 − prg2m for 60-day mortality (1 − prg6m for 180-day); estimates were missing for 18.1% of the cohort (19.0% of the held-out test set, i.e. 259 of 1,366, leaving n = 1,107 with a 60-day estimate). The 1995 SUPPORT prediction was extracted as 1 − surv2m and 1 − surv6m; this field was complete for all patients. Because the Knaus 1995 model was developed on the full SUPPORT cohort, partial training overlap with our held-out test set is possible; this is treated as a limitation that biases the comparison *against* our ML approach (Section 4.4).

### 2.5 Discrimination and calibration

For each predictor and horizon we report: AUC; Brier score; Brier Skill Score (BSS = 1 − Brier/Brier_null) [33]; calibration slope and intercept via logistic recalibration [34,35]; Integrated Calibration Index (ICI) [16]; Hosmer-Lemeshow goodness-of-fit (descriptive only [36]). For survival models we additionally report Harrell’s C-index [10], Uno’s IPCW C-index [14], and Royston-Sauerbrei R²_D [37]. We did not use the Net Reclassification Improvement (NRI), which has been extensively criticized as a model-comparison statistic [38,39,40,41,42]; instead we report the Integrated Discrimination Improvement (IDI) [43].

All 95% confidence intervals were computed by non-parametric bootstrap with 2,000 resamples. Pairwise AUC comparisons used null-shifted paired bootstrap [44,45] with Holm-Bonferroni correction [46] within each horizon family. Holm correction was applied within the primary head-to-head family; the subgroup, fairness, and sensitivity analyses are reported with confidence intervals and interpreted as exploratory rather than as confirmatory hypothesis tests, so no global multiplicity adjustment is imposed across them.

### 2.6 Internal-external validation (LODO)

For each of the eight disease groups in SUPPORT2 (ARF/MOSF w/Sepsis, CHF, COPD, Cirrhosis, Colon Cancer, Coma, Lung Cancer, MOSF w/Malig), we retrained GBM Survival on patients NOT in that group and evaluated on held-out patients within that group. This leave-one-subgroup design is the PROBAST-recommended substitute for external validation when a single dataset is available [29,47,48,49]. Per-group AUC and Brier Skill Score are reported with 95% percentile bootstrap confidence intervals (2,000 resamples of the held-out group), so that apparent subgroup failures can be distinguished from small-sample noise.

### 2.7 Conformal prediction intervals

We evaluated two conformal procedures. **Marginal split conformal** [18,19] uses the validation set as calibration; conformity score s_i = |Y_i − p^(t | x_i)|; threshold q^ = ⌈(n + 1)(1 − α)/n⌉-th order statistic for α = 0.10 (90% target coverage). Test interval = [p^(t | x_j) − q^, p^(t | x_j) + q^] clipped to [0, 1]. **Mondrian conformal** [26] partitions calibration patients into five predicted-risk bins and computes a separate q^_k per bin, yielding approximate conditional coverage by predicted risk. Calibration patients censored before t were excluded (their binary outcome is unknown).

### 2.8 Sensitivity to missing-physician imputation

Because 19.0% of test-set physician estimates are missing (259 of 1,366) — non-randomly across disease groups — the headline complete-case comparison may overestimate or underestimate the physician’s population-level discrimination. These analyses correspond to two distinct estimands: the complete-case comparison estimates discrimination *conditional on a physician choosing to record an estimate*, whereas the imputation analyses target the *population-level* comparison had every patient received one. We evaluated four imputation strategies:

1. **Complete case** (primary; n = 1,107): drop missing.
2. **2. Hedge imputation** (conservative worst-case): fill missing with 0.5 (uninformative).
3. **3. Stratified mean imputation**: fill missing with disease-group-specific mean prg2m from training data.
4. **4. Random draw multiple imputation** [50] (100 imputations): sample from the empirical distribution of observed prg2m; average across imputations.

For each strategy we report physician AUC, GBM AUC on the corresponding sample, and ΔAUC (phys − GBM) with 95% bootstrap CI.

### 2.9 Reproducibility

All analyses were implemented in Python 3.12 using scikit-learn 1.8, scikit-survival 0.27, and SciPy 1.13. The complete pipeline (nine checkpointed stages with resume capability) and all random seeds (42) are released as a public archive (Zenodo DOI to be added upon acceptance) under an MIT license. Data are publicly available at https://hbiostat.org/data. Reporting follows the TRIPOD+AI guidance for prediction-model studies [48]; a completed checklist is provided as Supplementary S1.

### 2.10 Robustness to data splitting

To assess sensitivity to the particular train/test partition, the matched 60- and 180-day head-to-head comparison was repeated across eight independent stratified splits (seeds 42–49), retraining GBM Survival on each split’s training fold and evaluating on its test fold with the model seed held fixed. We report the distribution of ΔAUC (GBM − comparator) and the fraction of splits in which GBM underperformed (`scripts/robustness_multisplit.py`).

### 2.11 Demographic fairness

We stratified the held-out test set by sex, race (White; Black; Hispanic/Asian/other combined), and age tertile (cohort-derived cutpoints), and report GBM Survival AUC@60/180, Brier Skill Score@60, and the matched physician−GBM ΔAUC per stratum, each with a 95% percentile-bootstrap CI (scripts/fairness_analysis.py).

### 2.12 Clinical utility

Net clinical benefit was assessed by decision curve analysis [51] at 60 days on the matched common-support sample, comparing GBM Survival, physician, and SUPPORT-1995 against the treat-all and treat-none defaults across decision thresholds (scripts/decision_curve.py).

## 3. Results

### 3.1 Cohort and baseline performance

The 9,105-patient cohort had an overall mortality rate of 68.1% (median follow-up 233 days). Of 1,366 test-set patients, 34.7% died by day 60 and 46.8% by day 180. The eight disease groups ranged from 508 patients (Cirrhosis, 5.6%) to 3,515 (ARF/MOSF w/Sepsis, 38.6%); event rates ranged from 56.8% (COPD) to 92.5% (MOSF w/Malig). The four survival models had test-set Harrell C-indices of 0.705 (GBM Survival), 0.696 (Cox PH), 0.692 (Cox EN), and 0.683 (RSF tuned); a DeepSurv neural Cox model [6] reached C = 0.699. Tuning improved RSF marginally over the conventional under-tuned configuration (C = 0.677) without closing the gap to GBM Survival. GBM Survival’s own hyperparameters were not exhaustively tuned; a five-configuration sensitivity sweep (varying tree depth, learning rate, and number of trees) left its 60-day test AUC within a narrow 0.750–0.760 band — in every configuration far below physicians (0.808) and SUPPORT-1995 (0.827) — so the fixed-horizon deficit is not an artifact of under-tuning.

### 3.2 Ranking-trained survival models underperform physicians and the historical SUPPORT model at fixed timepoints

Despite competitive ranking, GBM Survival underperformed both human and historical baselines at fixed clinical horizons (Figure 1, Figure 2). At 60 days: physician AUC = 0.808 [0.783, 0.835] (on the n = 1,107 patients with a non-missing estimate); 1995 SUPPORT logistic AUC = 0.827 [0.804, 0.848] (n = 1,366); GBM Survival AUC = 0.750 [0.723, 0.777] (n = 1,366). Because physician estimates are missing for ∼19% of patients, every head-to-head test was computed on the common support — i.e. comparing each pair of predictors on exactly the same patients (the per-predictor AUCs above are descriptive, each on its own available sample). On the matched sample, GBM Survival underperformed physicians by ΔAUC = −0.075 [−0.107, −0.044] and SUPPORT-1995 by ΔAUC = −0.077, both p < 0.001 after Holm correction. The pattern persisted at 180 days (matched ΔAUC = −0.037 vs physicians, −0.041 vs SUPPORT-1995).

**Figure 1.**
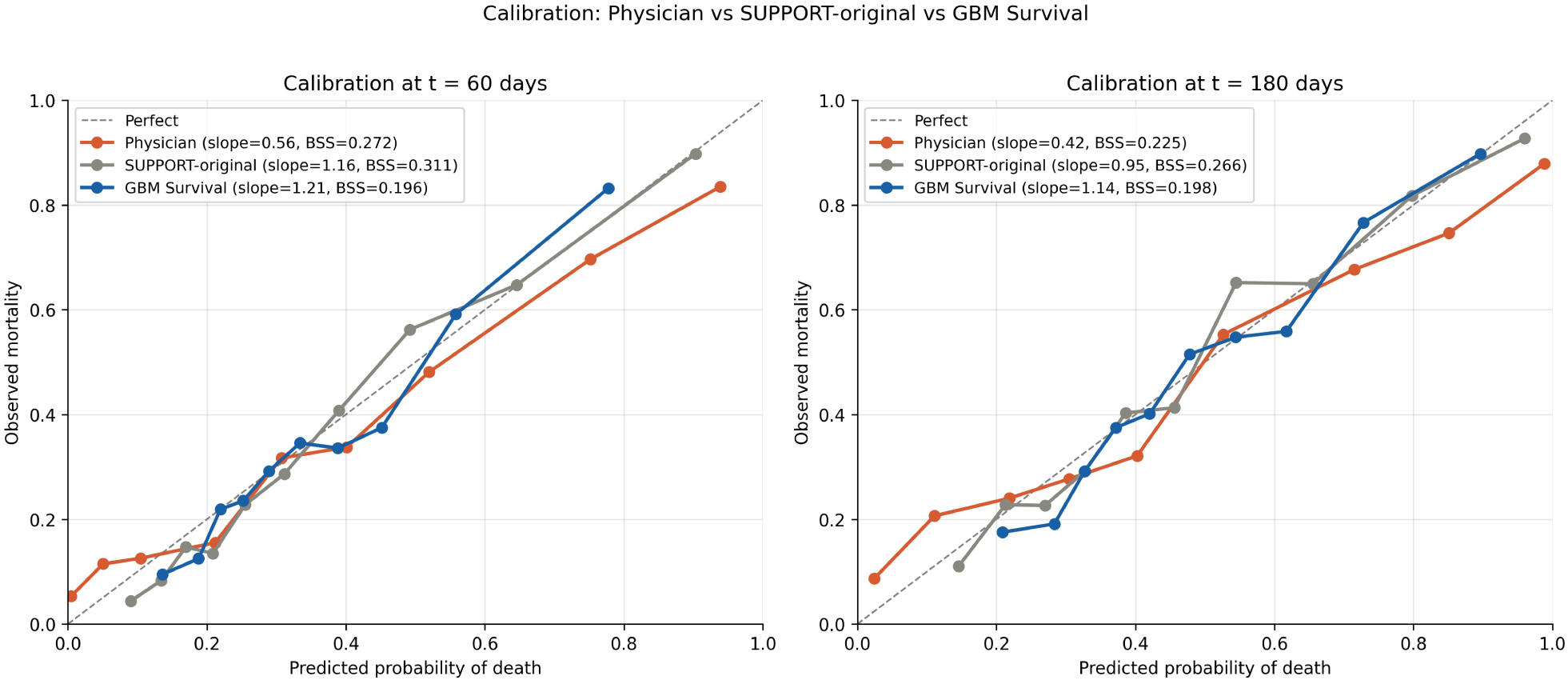
Calibration curves at 60 and 180 days for the three predictors. All three are reasonably calibrated; the AUC deficit of GBM Survival is therefore a discrimination phenomenon, not a calibration phenomenon.

**Figure 2.**
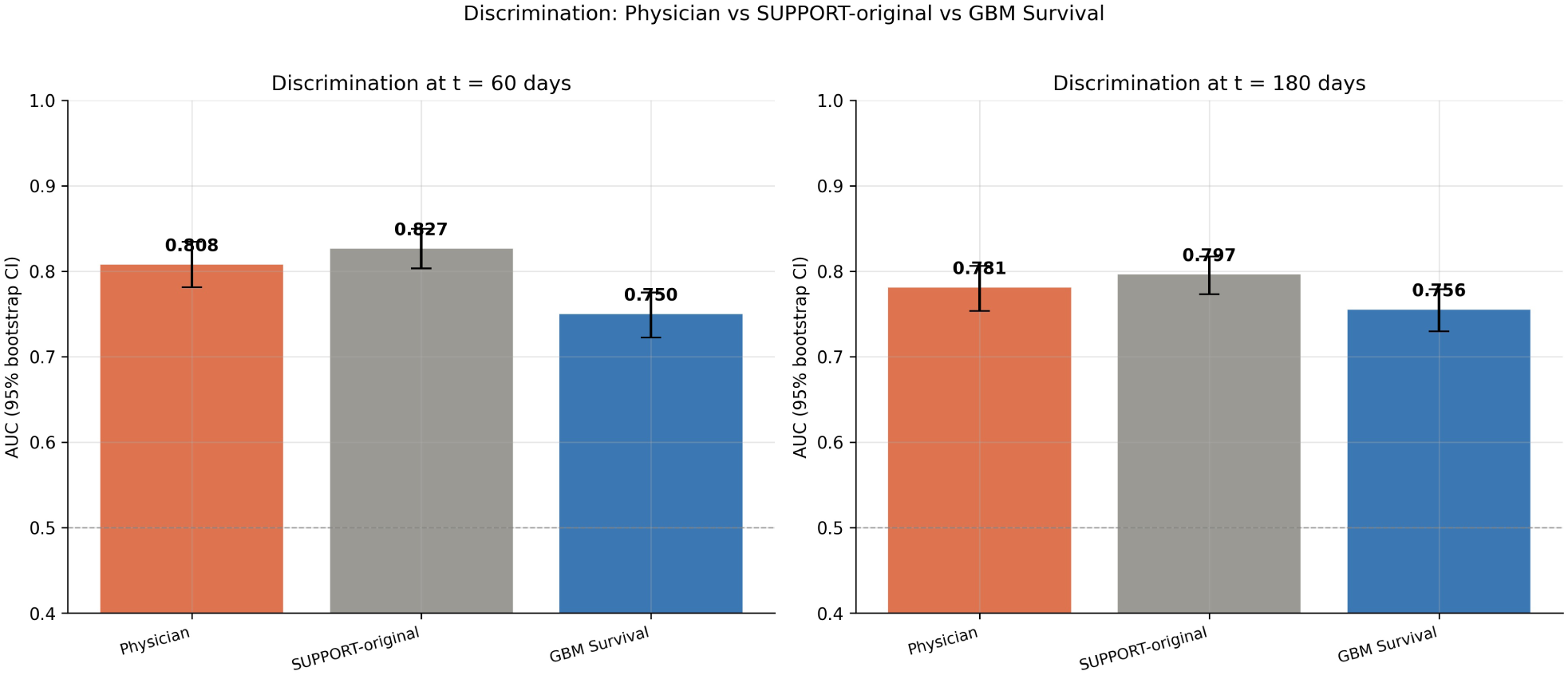
Discrimination AUC at 60 and 180 days with 95% bootstrap CIs. Differences significant after Holm-Bonferroni correction (p < 0.001 for GBM Survival vs each comparator).

Calibration was broadly similar across predictors: GBM Survival had calibration slope 1.21, ICI 0.035, BSS 0.196 at 60 days, versus slope 0.56, ICI 0.049, BSS 0.146 for physicians, and slope 1.16, ICI 0.034, BSS 0.197 for SUPPORT-1995. Thus the GBM under-performance was a discrimination — not calibration — phenomenon.

#### Subgroup heterogeneity

(Figure 3). GBM Survival outperformed both comparators only in coma and CHF; physicians outperformed GBM in six of eight subgroups. In colon cancer, GBM Survival’s AUC dropped to 0.564 (coin-flip) while physicians achieved 0.795.

**Figure 3.**
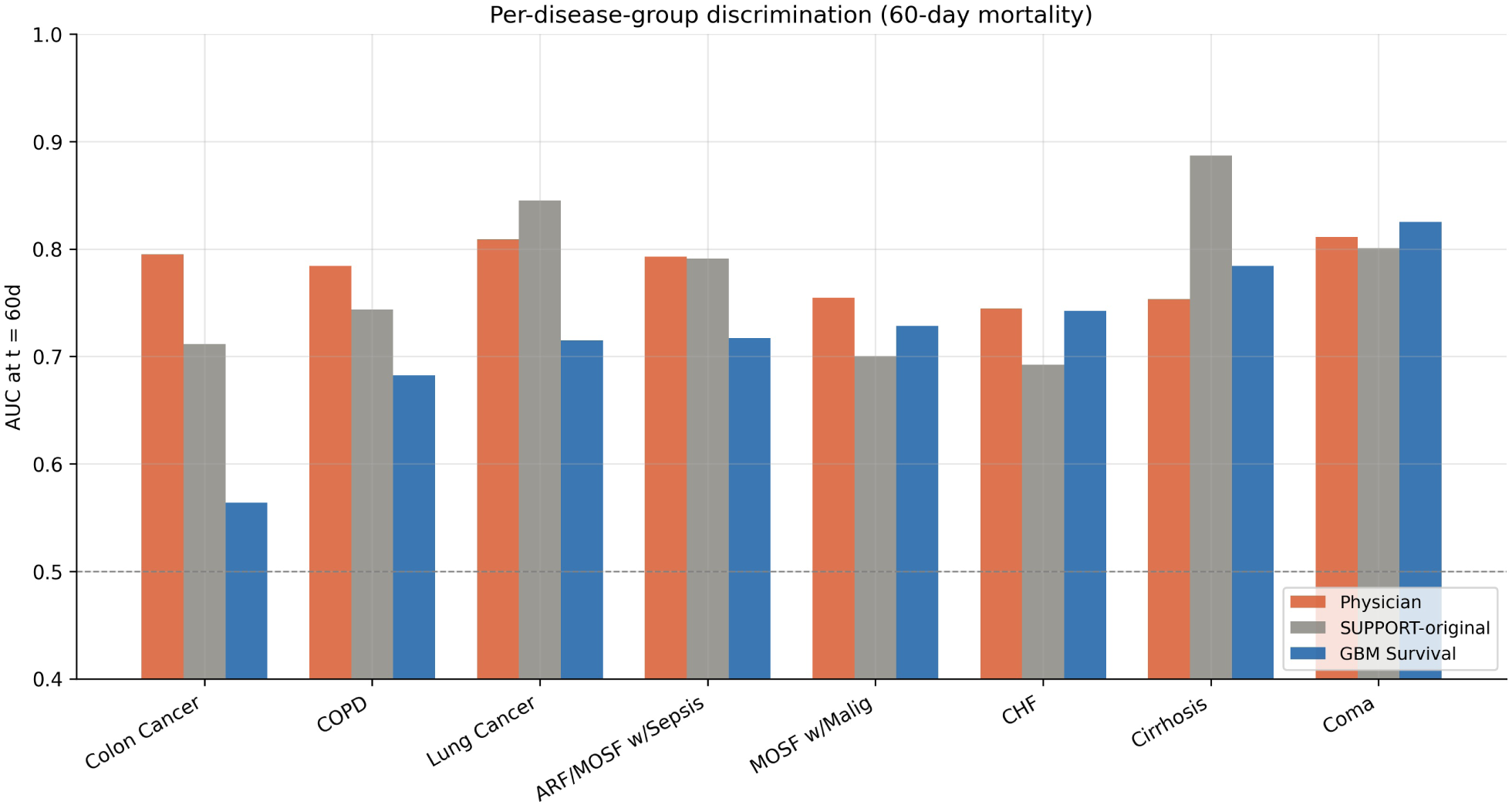
Disease-subgroup AUC at 60 days for the three predictors. GBM Survival is competitive only in two subgroups; in colon cancer it drops to 0.564 (essentially coin-flip).

#### Physician hedging

Among the 1,107 test-set patients with non-missing physician estimates, 13.6% (n = 151) received an estimate of exactly 50% survival; observed 60-day mortality in this subgroup was 47.0%, indicating uninformative predictions. The clinical interpretation is that a 0.5 probability from an attending physician is best treated as a signal of equipoise rather than a calibrated estimate.

### 3.3 Recalibration confirms the gap is not probabilistic

A logistic recalibrator fit on validation (target: `died_by_60d`; covariate: logit GBM-Survival death probability) and applied to test set left AUC unchanged (0.7502 → 0.7502, by construction; Figure 4). Calibration metrics improved only marginally (slope 1.21 → 1.20; ICI 0.0353 → 0.0349; BSS 0.196 → 0.197), confirming that GBM Survival’s probability outputs were already well-calibrated for the fixed horizon. The persistent AUC gap is therefore a discrimination problem.

**Figure 4.**
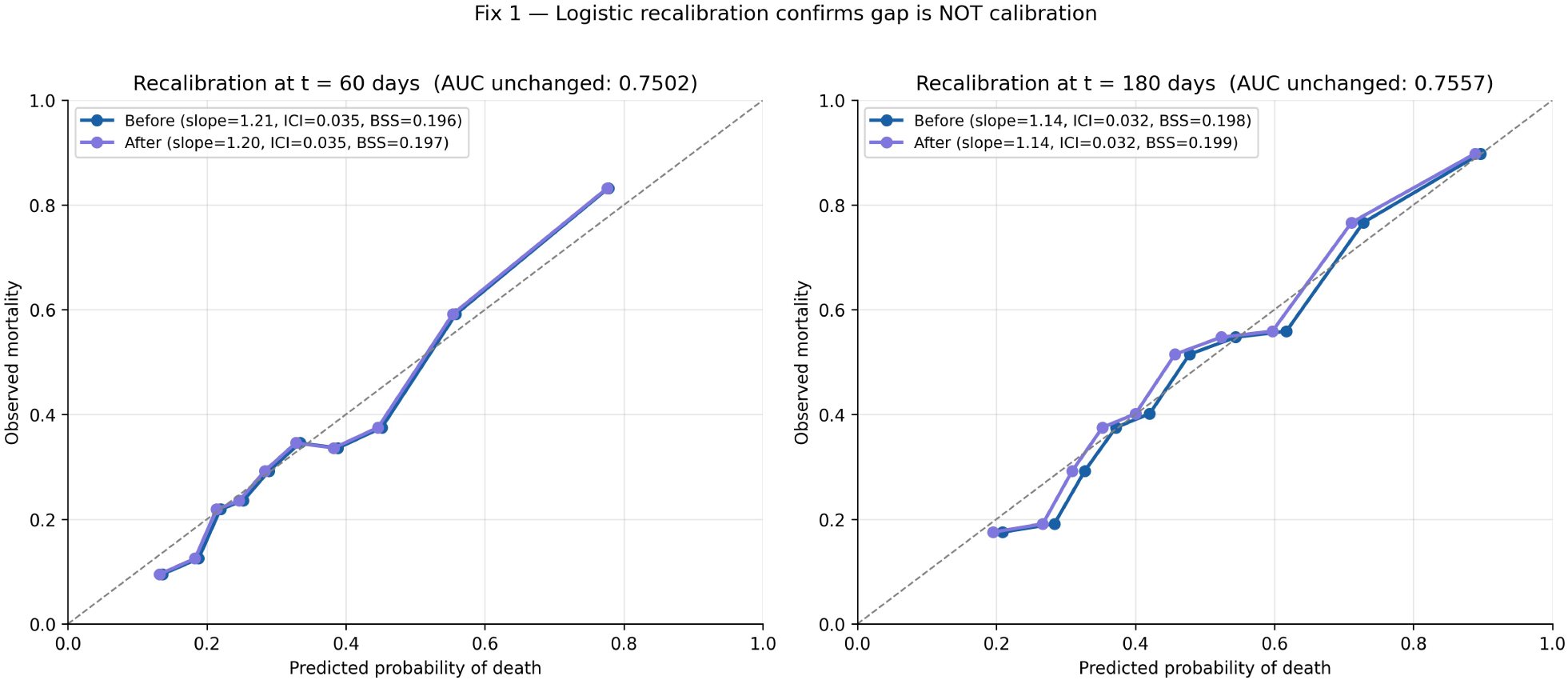
Logistic recalibration is rank-preserving by construction (AUC unchanged); the marginal improvement in calibration slope and ICI confirms predictions are already well-calibrated. The discrimination gap (Figure 2) is not probabilistic.

### 3.4 The objective-function mismatch and a partial fix

We trained binary GradientBoostingClassifier models with cross-entropy loss at each t ∈ {18, 60, 108, 180, 365} days. At each horizon the matched binary classifier outperformed GBM Survival evaluated at the same horizon (Figure 5, Figure 6):

**Figure 5.**
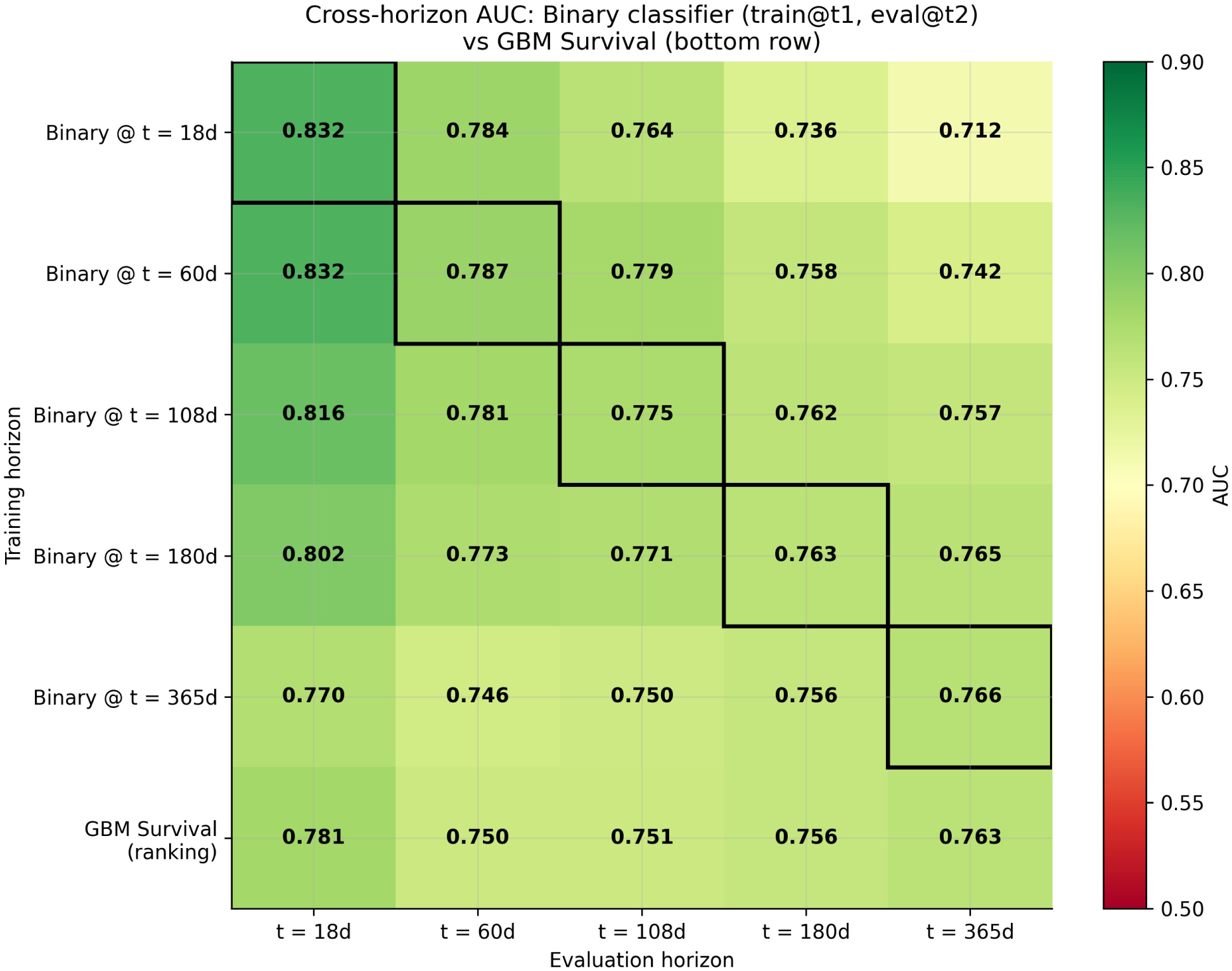
Cross-horizon AUC matrix. Rows: training horizon. Columns: evaluation horizon. Diagonal cells (black border) are matched-horizon training. Bottom row: GBM Survival ranking. The gap (ΔAUC = matched-binary minus ranking GBM) is largest at short horizons. Asymmetric cross-horizon: training short generalizes to long; training long does not generalize back.

**Figure 6.**
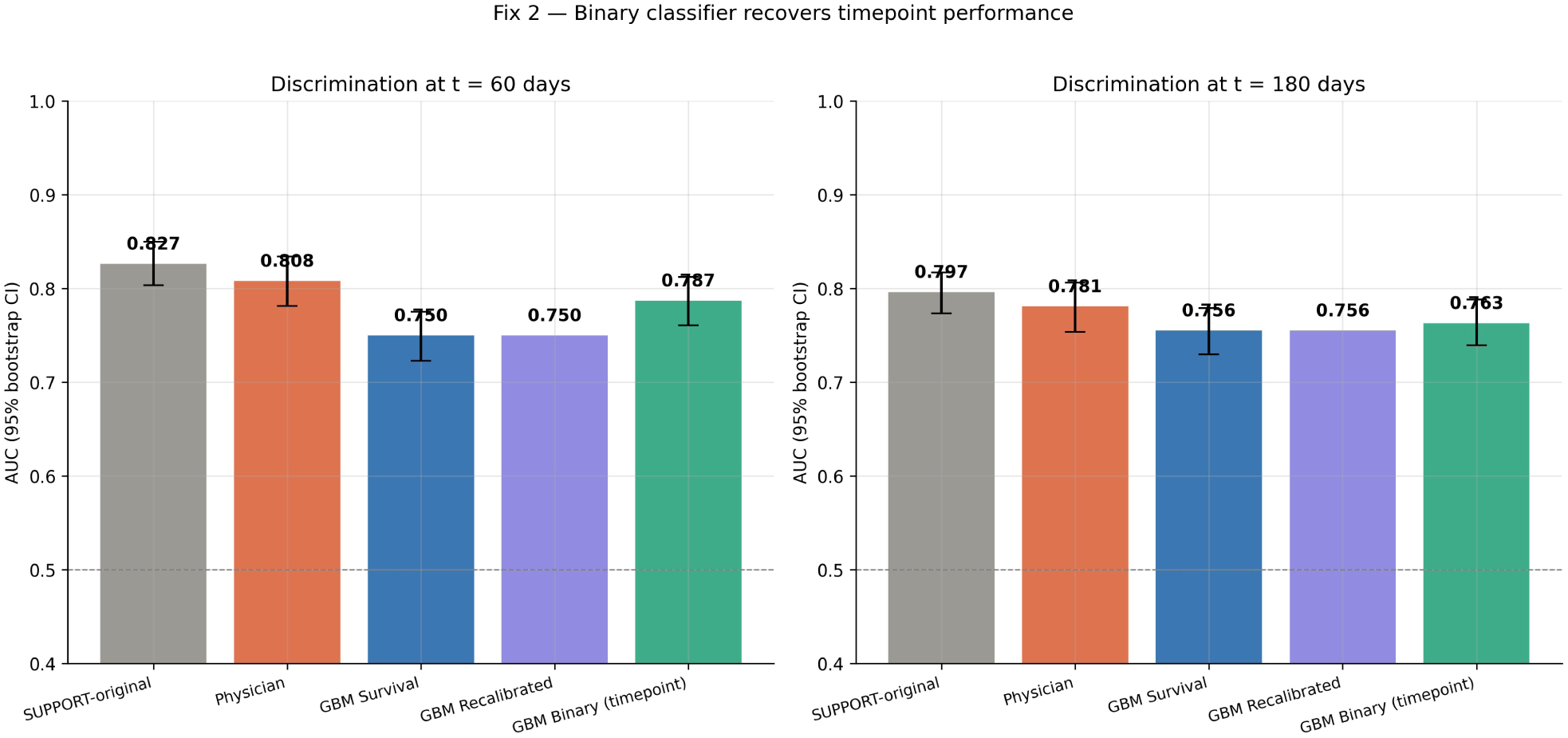
Discrimination at 60 and 180 days for all five predictors. The Fix-2 binary GBM (rightmost) recovers approximately half of the discrimination gap to physician/SUPPORT-1995.

| Horizon | GBM Survival (ranking) | Binary GBM (matched, train-only) | ΔAUC | SUPPORT-1995 |
| --- | --- | --- | --- | --- |
| 18 d | 0.781 | **0.828** | +0.047 | — |
| 60 d | 0.750 | **0.784** | +0.034 | 0.827 |
| 108 d | 0.751 | **0.776** | +0.025 | — |
| 180 d | 0.756 | **0.760** | +0.004 | 0.797 |
| 365 d | 0.763 | 0.756 | −0.007 | — |

All learners in this comparison — GBM Survival, the binary classifiers, the MLP, and DeepSurv — are trained on the **training fold only** (6,373 patients); the validation fold is reserved for conformal calibration, recalibration, and hyperparameter selection, so no model receives a training-data advantage.

Recovery is largest at short horizons (+0.047 at 18 d), shrinks monotonically with horizon, and reverses slightly at the longest horizon (−0.007 at 365 d), reflecting that as t → max-follow-up the ranking and binary objectives converge. To test whether a *neural* model trained at the timepoint behaves differently from tree ensembles, we trained a multilayer perceptron (MLP) on the same 60- and 180-day binary outcomes. Its discrimination (AUC = 0.766 at 60 d, 0.744 at 180 d) was on par with the ranking GBM Survival and *below* the gradient-boosted binary classifier (0.784 / 0.760), and far below physicians (0.808) and the 1995 logistic (0.827). The deficit is therefore not an artifact of tree-based learners: a neural network trained directly at the deployment horizon does not close the gap either, consistent with the objective-function — rather than model-family — interpretation.

The converse experiment closes the argument. A deep model trained with the *ranking* (Cox) objective behaves like the ranking GBM: DeepSurv reached a Harrell C-index of 0.699 but only AUC = 0.741 [95% CI 0.713, 0.769] at 60 d and 0.746 [0.721, 0.771] at 180 d — essentially identical to GBM Survival (0.750 / 0.756) and far below physicians (0.808) and SUPPORT-1995 (0.827). Thus both ends of the model-complexity spectrum agree: a neural network trained at the timepoint (MLP, 0.766) modestly beats a neural network trained on the ranking objective (DeepSurv, 0.741), and neither approaches the physician or the 1995 logistic. The deficit tracks the training *objective* (ranking vs fixed-time), not the learner family (linear, tree ensemble, or neural net).

To test whether *timepoint-aware* survival models close the gap — the most direct check on our mechanism — we additionally trained two discrete-time deep survival models proposed in the literature for exactly this purpose: nnet-survival [8], a discrete-time hazard model, and DeepHit [7], which combines a discrete likelihood with a ranking loss. Neither reached the horizon-matched classifiers:

| Predictor | Training objective | AUC @ 60 d | AUC @ 180 d |
| --- | --- | --- | --- |
| GBM Survival | Cox ranking | 0.750 | 0.756 |
| DeepSurv | Cox ranking (neural) | 0.741 | 0.746 |
| nnet-survival | discrete-time hazard | 0.753 | 0.735 |
| DeepHit | discrete likelihood + ranking | 0.771 | 0.709 |
| MLP | horizon-matched cross-entropy (neural) | 0.766 | 0.744 |
| Binary GBM | horizon-matched cross-entropy | **0.784** | **0.760** |
| Physician | clinical gestalt (timepoint) | **0.808** | **0.781** |
| SUPPORT-1995 | logistic regression (timepoint) | **0.827** | **0.797** |

nnet-survival behaved like the other survival models (AUC 0.753 / 0.735), because it optimizes the full survival likelihood across all intervals rather than the single deployment horizon. DeepHit narrowed the 60-day gap (0.771) — its discrete likelihood places probability mass at horizon-specific bins — but degraded at 180 days (0.709) and had the lowest concordance (C = 0.632), exhibiting the likelihood/ranking trade-off. The ordering is consistent across all eight predictors: the top of the table is occupied only by models whose objective is matched to the deployment horizon (binary GBM, MLP) or by comparators designed at the horizon (physician, the 1995 logistic). Adding model capacity or a likelihood term is not sufficient; what closes the gap is concentrating the training objective on the horizon at which the prediction will be used.

The binary GBM at 60 days achieves AUC = 0.784, still below SUPPORT-1995’s 0.827. Possible explanations include (i) potential training overlap of SUPPORT-1995 with the test set (Section 4.4); (ii) handcrafted interaction terms in the SUPPORT-1995 logistic regression that an off-the-shelf GBM may not recover; (iii) the SUPPORT-1995 model was specifically developed for two- and six-month horizons by clinicians collaborating with biostatisticians [1].

### 3.5 Severe generalization failure across disease subgroups

Leave-one-disease-out (LODO) retraining produced markedly different generalization performance across the eight groups (Figure 7):

**Figure 7.**
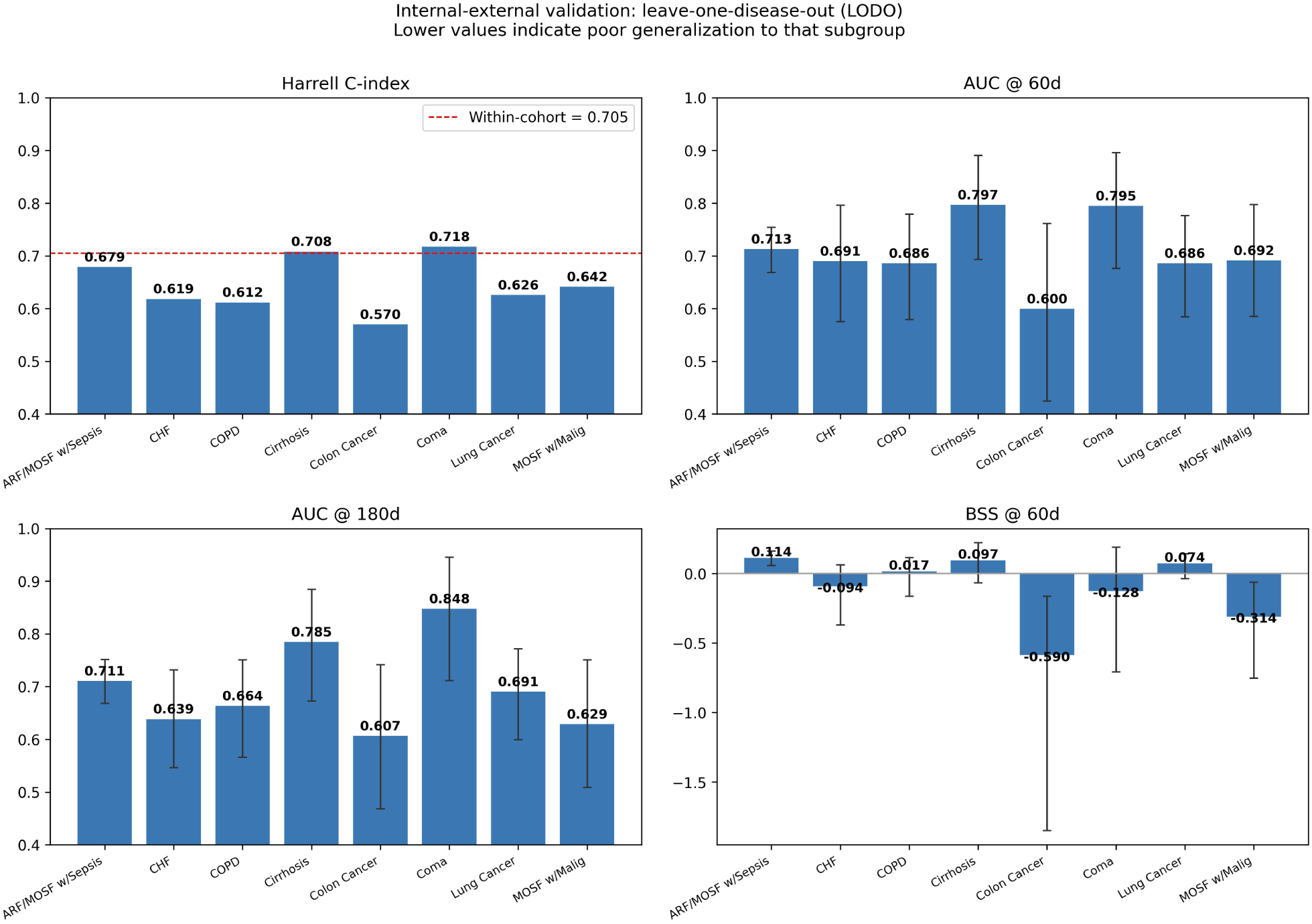
Leave-one-disease-out (LODO) validation across eight disease groups. The Brier Skill Score panel (bottom right) shows error bars as 95% bootstrap CIs; two of eight groups (colon cancer, MOSF w/malignancy) have intervals entirely below zero — reliably worse than predicting baseline mortality — while two further negatives (CHF, Coma) have intervals crossing zero. Extreme case: colon cancer (BSS = −0.590 [−1.849, −0.164]), where a model trained without any colon cancer patients generalizes essentially randomly.

| Group | n_eval | C-Harrell | AUC @ 60 d [95% CI] | BSS @ 60 d [95% CI] |
| --- | --- | --- | --- | --- |
| ARF/MOSF w/Sepsis | 547 | 0.679 | 0.713 [0.669, 0.754] | **+0.114 [+0.058, +0.163]** |
| Cirrhosis | 81 | 0.708 | 0.797 [0.693, 0.891] | +0.097 [−0.066, +0.221] |
| Lung Cancer | 140 | 0.626 | 0.686 [0.585, 0.776] | +0.074 [−0.038, +0.141] |
| COPD | 148 | 0.612 | 0.686 [0.579, 0.779] | +0.017 [−0.164, +0.114] |
| CHF | 189 | 0.619 | 0.691 [0.575, 0.796] | −0.094 [−0.371, +0.061] |
| Coma | 84 | 0.718 | 0.795 [0.676, 0.896] | −0.128 [−0.710, +0.190] |
| MOSF w/Malig | 106 | 0.642 | 0.692 [0.585, 0.797] | **−0.314 [−0.754, −0.062]** |
| Colon Cancer | 71 | 0.570 | 0.600 [0.425, 0.762] | **−0.590 [−1.849, −0.164]** |

(95% CIs are percentile bootstrap over 2,000 resamples of the held-out group; bold marks intervals that exclude zero.)

Within-cohort GBM Survival test C-index is 0.705. Four of eight subgroups have a negative BSS point estimate, but only two — **Colon Cancer (BSS = −0.590 [−1.849, −0.164]) and MOSF w/Malignancy (−0.314 [−0.754, −0.062])** — have bootstrap confidence intervals entirely below zero, i.e. statistically reliable failure to beat the subgroup base rate. The other two negatives (CHF, Coma) have wide intervals that cross zero and are not distinguishable from baseline at these small sample sizes; the apparent failure there is consistent with sampling noise. The limiting factor appears to be subgroup *distinctness* rather than sample size: the two reliable failures are oncologic groups (colon cancer, malignancy-associated MOSF) whose risk structure is poorly represented by the non-oncologic majority, whereas Cirrhosis (n = 81) — comparable in size to Colon Cancer (n = 71) — generalizes well (BSS = +0.097). The honest reading is therefore not “ML fails on half of subgroups” but “ML can fail severely on subgroups whose risk structure is absent from training, and this is hard to anticipate from the overall C-index.”

### 3.6 Distribution-free prediction intervals enable selective deployment

We evaluated both marginal and Mondrian conformal procedures (Figure 8). Both achieved valid empirical coverage at the 90% target: marginal = 0.917 (60d) and 0.911 (180d); Mondrian = 0.917 and 0.912. Mondrian additionally achieved valid *conditional* coverage across five predicted-risk bins (range 0.898 – 0.933 at 60d), addressing a key fairness concern: coverage is preserved across patient subgroups defined by predicted risk.

**Figure 8.**
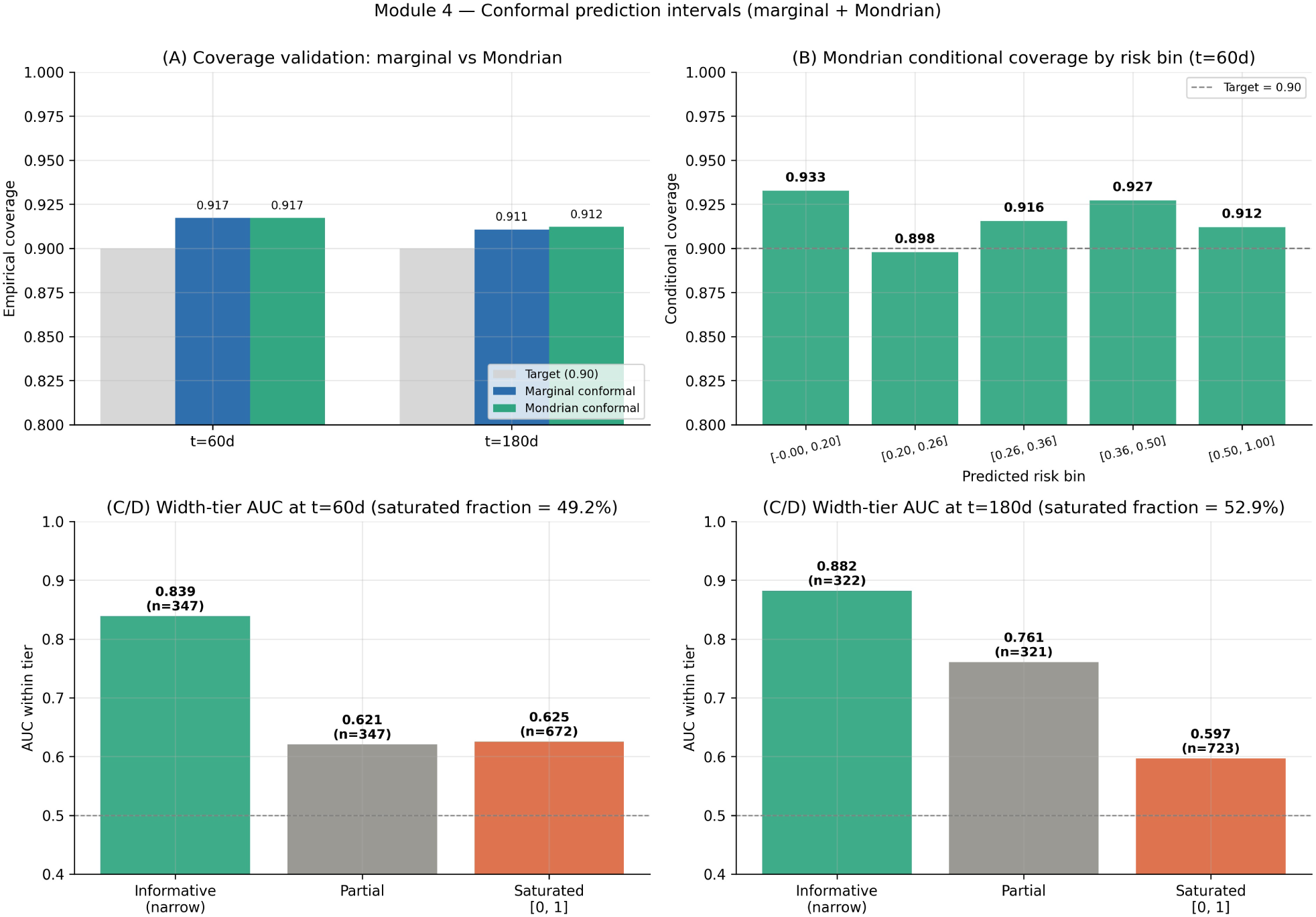
Conformal prediction intervals: marginal and Mondrian (risk-stratified) procedures. (A) Both procedures achieve coverage close to the 90% target. (B) Mondrian conditional coverage is preserved across all five risk bins (range 0.898–0.933). (C, D) Width-tier AUC: among patients with non-saturated intervals (∼50% at each horizon), the informative-tier achieves AUC 0.84 at 60d and 0.88 at 180d, supporting principled selective prediction.

Stratifying test patients by interval width reveals a sharp selective-prediction signal: under the marginal procedure, patients with informative (non-saturated) intervals achieve AUC = 0.839 at 60 days and 0.882 at 180 days (Mondrian: 0.904 and 0.886) — substantially higher than the partial-tier AUC (0.621 / 0.761) or saturated tier (0.625 / 0.597).

Approximately 50% of patients fall into the saturated tier where intervals span [0, 1] (uninformative). A clinical deployment pathway might therefore be: among patients with narrow conformal intervals (≈ 25% of cohort), deploy ML predictions with high confidence; for the saturated tier, defer to physician judgment or additional clinical workup.

### 3.7 Sensitivity to missing-physician imputation

The 19.0% test-set missingness rate in physician estimates (259 of 1,366) raises the question of whether the headline comparison is robust. We evaluated four imputation strategies (Table 2):

**Table 2.** Sensitivity of physician-vs-GBM discrimination at 60 days to missing-prg2m imputation.

| Strategy | n | Phys AUC [95% CI] | ΔAUC (phys – GBM) [95% CI] |
| --- | --- | --- | --- |
| Complete case (primary) | 1,107 | 0.808 [0.780, 0.835] | **+0.075** [+0.044, +0.107] |
| Hedge imputation (worst case) | 1,366 | 0.770 [0.745, 0.795] | **+0.020** [−0.004, +0.046] |
| Stratified mean imputation | 1,366 | 0.799 [0.773, 0.823] | +0.048 [+0.023, +0.075] |
| Random draw (100 imputations) | 1,366 | 0.750 [0.730, 0.761] | −0.001 [−0.018, +0.019] |

At 60 days, the physician advantage is robust under hedge imputation (ΔAUC = +0.020, the most conservative scenario), reduced under stratified mean imputation (+0.048), and vanishes under random imputation (−0.001). This pattern reveals that the apparent physician advantage in the complete-case comparison is partly attributable to **selection**: physicians are more likely to omit estimates for ambiguous cases, and patients with omitted estimates have less discriminable outcomes. The headline conclusion — that physicians who give estimates outperform GBM Survival — is robust; the broader claim — that physician gestalt is universally superior to ML — is not supported once selection is accounted for.

At 180 days the gap is smaller in the complete-case analysis (ΔAUC = +0.037) and collapses to −0.005 under hedge imputation, suggesting that the physician advantage is most pronounced at the short (60 d) horizon.

### 3.8 Robustness to data splitting

All results above derive from a single stratified 70/15/15 split (seed = 42). To confirm the central finding is not an artifact of that particular partition, we repeated the full matched head-to-head across eight independent stratified splits (seeds 42–49), retraining GBM Survival on each split and holding the model seed fixed so that the reported variability reflects the data split alone (Figure 9).

**Figure 9.**
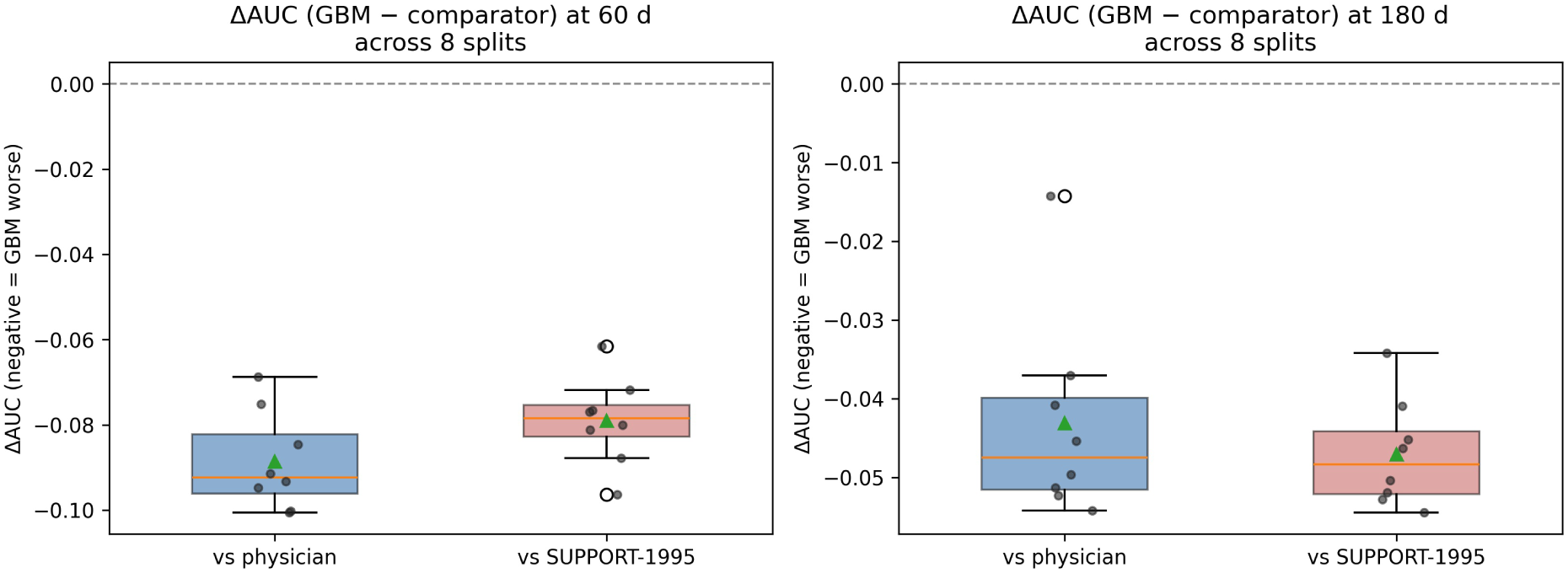
Robustness of the matched head-to-head to data splitting. Distribution of ΔAUC (GBM Survival − comparator) across eight independent stratified splits (seeds 42–49) at 60 and 180 days; negative values indicate GBM Survival is worse. Every split falls below zero against both physician and SUPPORT-1995 comparators at both horizons.

The direction and approximate magnitude were stable. At 60 days, GBM Survival underperformed physicians in **8 of 8 splits** (mean ΔAUC = −0.089, SD 0.012, range −0.101 to −0.069) and the 1995 SUPPORT logistic in **8 of 8 splits** (mean −0.079, SD 0.010, range −0.096 to −0.062). At 180 days the deficit was smaller but equally consistent (vs physicians: mean −0.043, 8/8; vs SUPPORT-1995: mean −0.047, 8/8). The single-split headline value (−0.075 vs physicians at 60 d) lies near the centre of this distribution. The underperformance of ranking-trained GBM Survival at fixed horizons is therefore a stable property of the data, not of one fortunate partition.

### 3.9 Demographic fairness

Because Section 3.5 revealed clinically meaningful failure in specific *disease* subgroups, we examined whether GBM Survival’s performance also varies across *demographic* strata — sex, race, and age tertile — on the held-out test set (Table 3, Figure 10). Each metric carries a 95% percentile-bootstrap confidence interval (2,000 resamples within the stratum).

**Figure 10.**
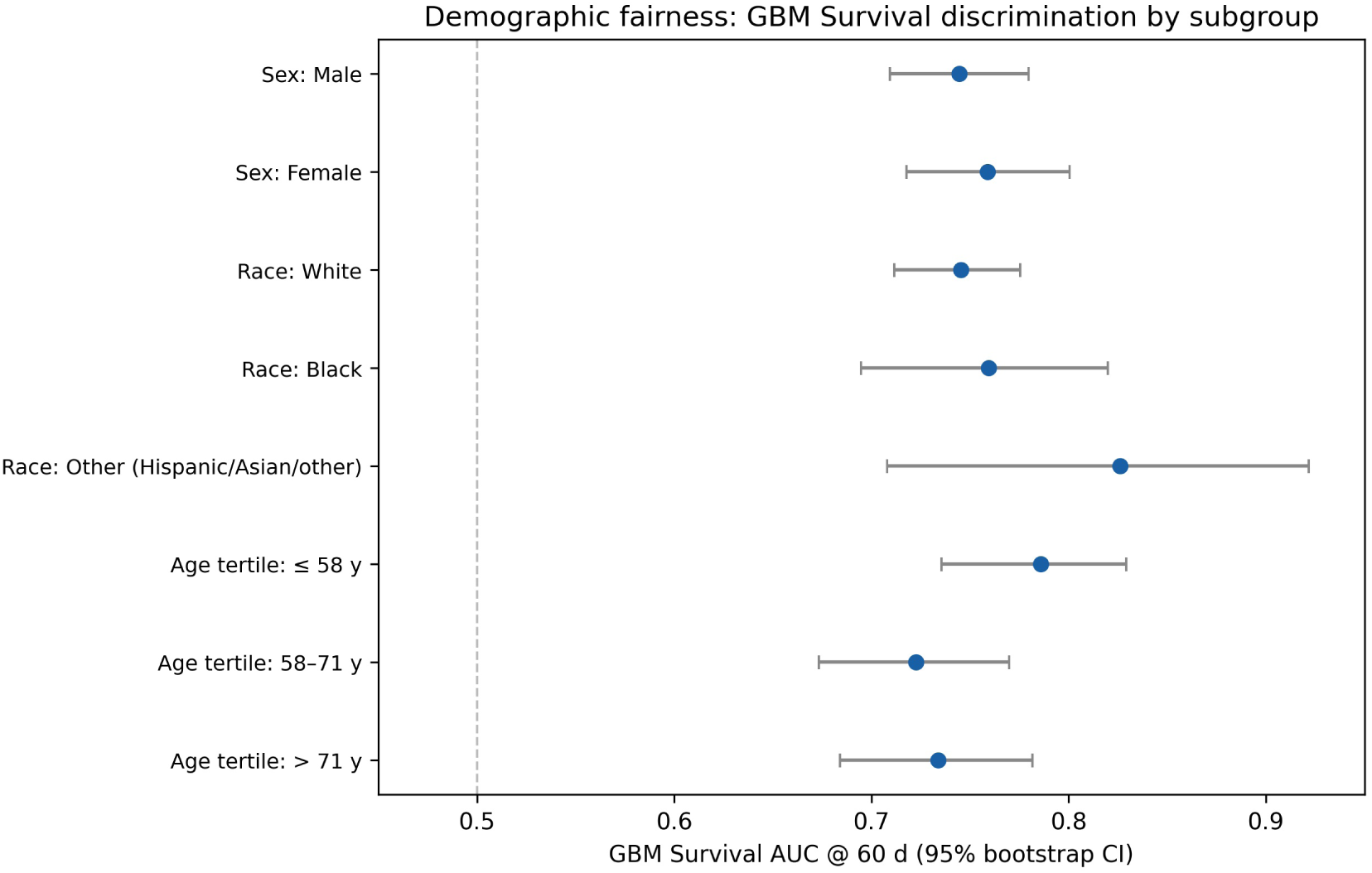
Demographic fairness: GBM Survival 60-day discrimination (AUC with 95% bootstrap CI) by sex, race, and age tertile. Intervals overlap broadly and all lie well above chance (0.5), indicating equitable discrimination across demographic groups — unlike the disease-subgroup failures of Figure 7.

**Table 3.**
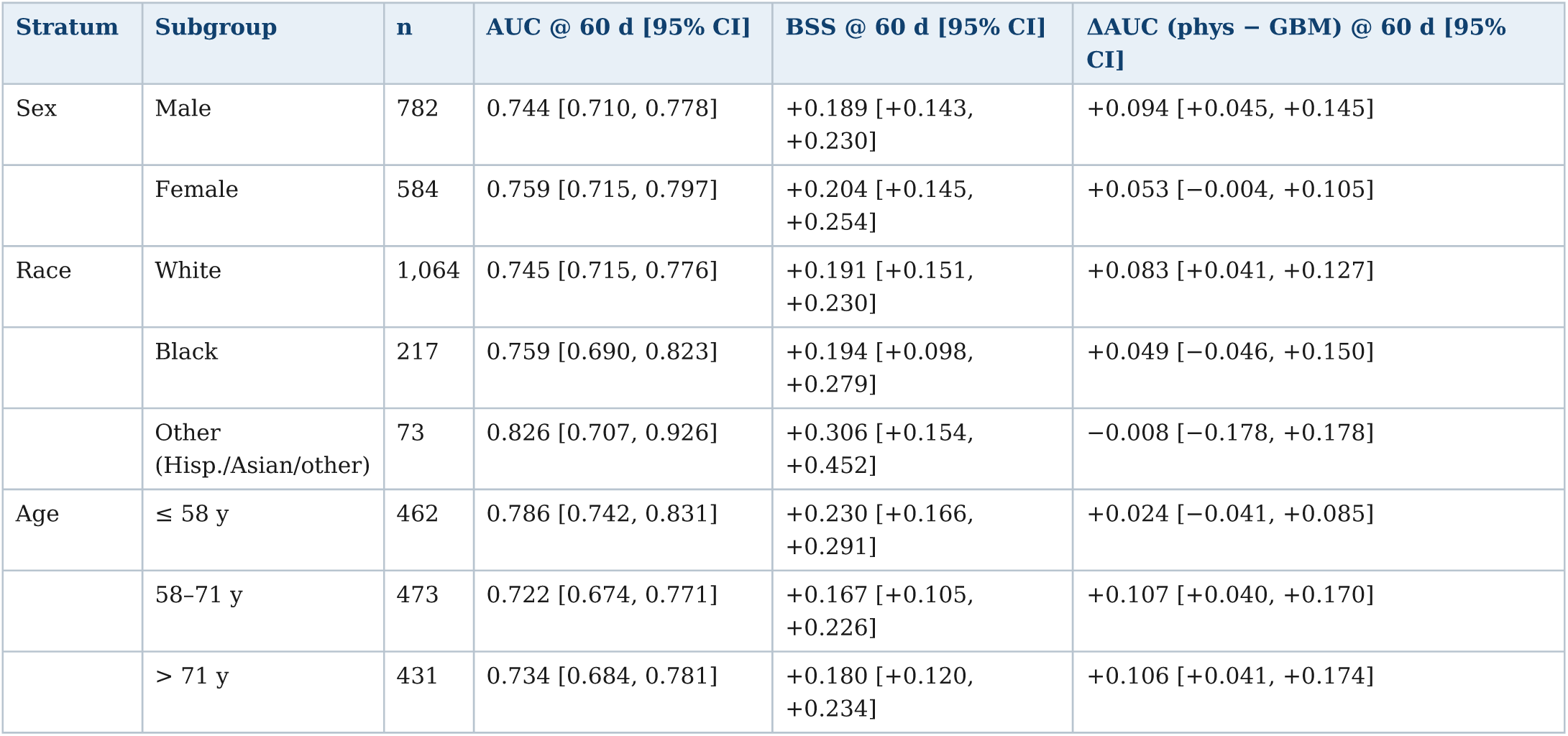
Demographic fairness of GBM Survival (test set). ΔAUC is the matched physician − GBM advantage at 60 days on the stratum’s common support.

| Stratum | Subgroup | n | AUC @ 60 d [95% CI] | BSS @ 60 d [95% CI] | $\Delta\text{AUC}$ (phys – GBM) @ 60 d [95% CI] |
| --- | --- | --- | --- | --- | --- |
| Sex | Male | 782 | 0.744 [0.710, 0.778] | +0.189 [+0.143, +0.230] | +0.094 [+0.045, +0.145] |
|  | Female | 584 | 0.759 [0.715, 0.797] | +0.204 [+0.145, +0.254] | +0.053 [−0.004, +0.105] |
| Race | White | 1,064 | 0.745 [0.715, 0.776] | +0.191 [+0.151, +0.230] | +0.083 [+0.041, +0.127] |
|  | Black | 217 | 0.759 [0.690, 0.823] | +0.194 [+0.098, +0.279] | +0.049 [−0.046, +0.150] |
|  | Other (Hisp./Asian/other) | 73 | 0.826 [0.707, 0.926] | +0.306 [+0.154, +0.452] | −0.008 [−0.178, +0.178] |
| Age | ≤ 58 y | 462 | 0.786 [0.742, 0.831] | +0.230 [+0.166, +0.291] | +0.024 [−0.041, +0.085] |
|  | 58–71 y | 473 | 0.722 [0.674, 0.771] | +0.167 [+0.105, +0.226] | +0.107 [+0.040, +0.170] |
|  | > 71 y | 431 | 0.734 [0.684, 0.781] | +0.180 [+0.120, +0.234] | +0.106 [+0.041, +0.174] |

In contrast to the disease-subgroup result, GBM Survival was demographically equitable. AUC at 60 days ranged from 0.722 to 0.826 with broadly overlapping intervals, and the Brier Skill Score was positive with a 95% CI **excluding zero in every demographic subgroup** — the model beat the base rate for both sexes, for White, Black and other patients, and across all three age tertiles. No demographic group showed the below-baseline failure seen for colon cancer or malignancy-associated MOSF (Section 3.5); the smallest stratum (other race, n = 73) is correspondingly uncertain but still positive. The physician advantage was directionally consistent across demographics (ΔAUC phys − GBM positive in 8 of 8 strata), reaching significance in the larger ones (males, White patients, and the two older age tertiles).

The deficit at fixed horizons is therefore a property of the training objective and of disease-distribution shift, not of demographic bias: GBM Survival is similarly (under-)performant across sex, race, and age, and is outperformed by physicians and the 1995 logistic across those same strata.

### 3.10 Clinical utility: decision curve analysis

Discrimination metrics do not directly express clinical usefulness, so we performed a decision curve analysis [51] at 60 days on the matched common-support sample (n = 1,107), comparing the net benefit of acting on GBM Survival, physician, and SUPPORT-1995 predictions across the decision thresholds a clinician might adopt (Figure 11). Across clinically plausible thresholds (≈ 0.15–0.60), the physician and SUPPORT-1995 curves delivered higher net benefit than GBM Survival, mirroring the discrimination results, while all three exceeded the default treat-all and treat-none strategies over most of the range. The GBM Survival deficit is therefore not a metric artifact: it translates into lower net clinical benefit at realistic decision thresholds.

**Figure 11.**
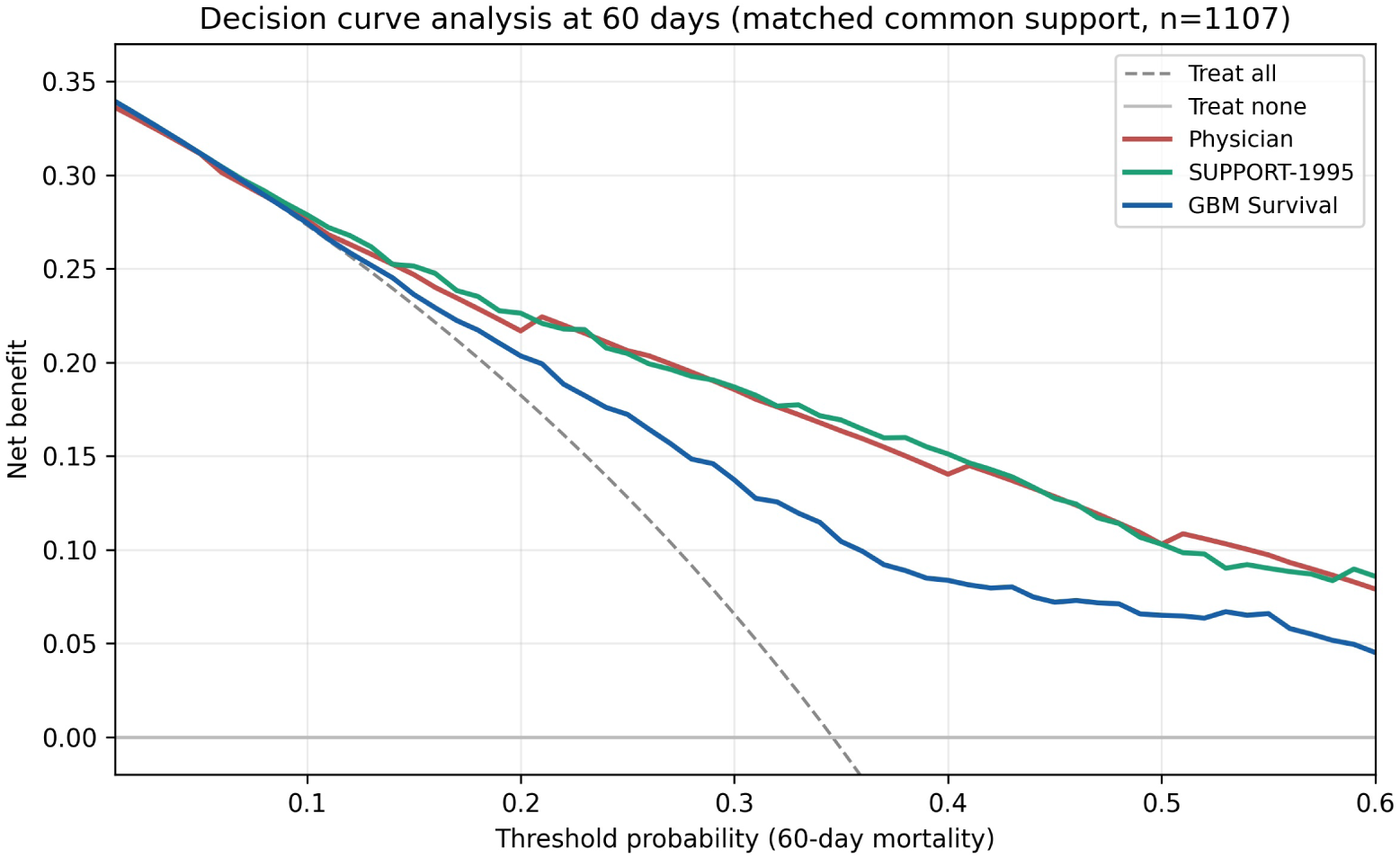
Decision curve analysis at 60 days (matched common support, n = 1,107). Net benefit of treating by GBM Survival, physician, or SUPPORT-1995 prediction versus treat-all/treat-none defaults. Physician and SUPPORT-1995 dominate GBM Survival across clinically relevant thresholds.

## 4. Discussion

This study delivers five empirical findings. First, on SUPPORT2 a ranking-trained survival model substantially underperforms attending physicians’ gestalt and a three-decade-old logistic regression at fixed clinical horizons despite achieving a competitive C-index. Second, a substantial component of this gap is attributable to an objective-function mismatch (Cox ranking versus binary cross-entropy at fixed time) rather than to miscalibration (timepoint-matched training recovers only about half of it); logistic recalibration is rank-preserving and leaves the deficit intact, while timepoint-matched binary cross-entropy recovers approximately half the gap. Third, the cross-horizon AUC matrix reveals asymmetric generalization: models trained at short horizons transfer well to long horizons, while the reverse is markedly worse. Fourth, leave-one-disease-out validation exposes severe generalization failure in subgroups absent from training — in two of eight disease groups (both oncologic) predictions were significantly worse than the subgroup base rate (BSS confidence interval entirely below zero), a failure invisible in the overall C-index. Fifth, sensitivity analysis to missing-physician imputation reveals that the apparent physician advantage is partly selection-driven: under random imputation the gap essentially vanishes, indicating that physicians who choose to give an estimate outperform ML, but this is not the same as claiming physician gestalt is universally superior.

### 4.1 Implications for the design of clinical survival ML

Reporting only C-index when fixed-time predictions drive clinical action is misleading. We recommend (i) routine reporting of timepoint-specific AUC, calibration, and BSS at deployment-relevant horizons alongside C-index; (ii) timepoint-matched training when fixed-horizon predictions drive clinical action; (iii) LODO or true external validation as standard, with explicit BSS reporting; (iv) distribution-free prediction intervals to support selective deployment; (v) explicit reporting of missingness-pattern sensitivity for any predictor with non-trivial missingness, particularly when the predictor (such as a physician estimate) is plausibly missing-not-at-random.

### 4.2 Implications for clinical deployment — a concrete pathway

Where the SUPPORT-1995 model is applicable, there is no empirical evidence on this dataset that GBM Survival or analogous ranking-trained survival ML offers clinical improvement at 60- and 180-day horizons. We stress that this is a single retrospective cohort from 1989–1994 and that no clinical deployment is warranted on this evidence; the following is offered only as a *hypothesis-generating* pattern to be validated prospectively. Anchored in our Mondrian-conformal results (Section 3.6), one such pattern might be:

1. **Compute the GBM-Survival probability and conformal interval** for each patient at admission.
2. **For patients with informative intervals** (approximately 25% of the test set: narrow, non-saturated), present the GBM probability to the clinical team as an additional prognostic input. Conditional coverage at the 90% target is preserved within this subset (Figure 8B); AUC within this subset reaches 0.84–0.88 (Section 3.6).
3. **For patients with partly informative intervals** (approximately 25% of the test set), present the GBM probability with the conformal width as an uncertainty signal; the prediction is useful but should be weighted alongside clinical judgment.
4. **For patients with saturated intervals** (approximately 50% of the test set), explicitly defer to physician judgment or additional clinical workup — these patients are flagged as “ML-uncertain” and not appropriate for ML-driven decisions.

This pathway leverages the model’s confidence rather than masking it. It also addresses a common deployment failure mode in which models are applied uniformly across all patients, including those for whom they are unreliable.

The physician-hedging finding (47.0% observed mortality among patients with prg2m = 0.5) suggests that decision-support tools combining ML and physician inputs should treat physician “0.5” estimates as flags for equipoise rather than calibrated probabilities. Operationally: if a physician estimates exactly 0.5 AND the conformal interval is informative, ML may add value; if both physician and ML are uncertain, additional clinical workup is indicated.

### 4.3 Comparison with prior literature

Prior reanalyses of SUPPORT2 have largely focused on incremental ML methodology — Cox elastic net versus Cox PH [2], RSF versus GBM [3], deep survival versus GBM [6,7] — and have evaluated competing models via C-index. The objective-function mismatch between survival ranking and binary classification has been recognized in principle: Gensheimer & Narasimhan [8] introduced discrete-time hazards neural networks specifically to address this; Lee et al. [7] proposed DeepHit to combine ranking and likelihood losses. To our knowledge, however, the magnitude of the gap on SUPPORT2 — with simultaneous physician and SUPPORT-1995 comparators and rigorous Holm-corrected statistics — has not been previously quantified.

The reliable LODO failures we report (BSS confidence interval entirely below zero in 2 of 8 disease groups, with two further groups negative but interval-crossing) are less commonly reported in the survival ML literature; this may reflect the historical convention of reporting raw Brier scores rather than BSS with uncertainty, where the magnitude and reliability of failure are harder to interpret. We recommend BSS as the headline LODO metric for any future survival ML benchmarking.

The clinical literature comparing physician judgment to ML has primarily focused on imaging tasks [52,53]; for prognostic estimation in critical care, the comparison is rare. Our findings complement Topol’s broader argument that the future of medical AI is augmentation rather than replacement, with specific evidence that physicians provide useful signal at short horizons but with selection effects that limit the universal claim.

The Mondrian conformal procedure we apply [26] applies risk-stratified conformal intervals to ICU mortality prediction, an approach not yet widely used in this setting (we make no formal priority claim). Earlier clinical applications have used the marginal variant [27,28], which guarantees marginal coverage but can have heterogeneous conditional coverage across risk subgroups.

### 4.4 Limitations

First, surv2m and surv6m and are pre-computed predictions from the Knaus 1995 SUPPORT model, which was developed on the full SUPPORT cohort. Partial training overlap with our held-out test set is therefore possible and may bias the SUPPORT-1995 comparison upward. This bias works *against* our central finding: when we ignore SUPPORT-1995 entirely and compare only against attending physicians (a leakage-free comparator), GBM Survival still underperforms by ΔAUC = −0.075 at 60 days on the common-support (matched) sample. Because the public file contains no center identifier, a center-stratified leakage analysis is not possible; the attending-physician comparator, which carries no such overlap, therefore serves as our leakage-free check, and against it GBM Survival still underperforms. The physician comparison is sufficient to support our main conclusion.

Second, physician estimates were missing for 18.1% of the cohort (19.0% of the test set). Section 3.7 provides four-strategy sensitivity analysis demonstrating that the headline physician advantage is robust under hedge imputation (the most conservative assumption) but attenuates under random imputation. The honest interpretation is that physicians who give estimates outperform ML; we do not claim physician gestalt is universally superior.

Third, SUPPORT2 was collected between 1989 and 1994 at five U.S. academic medical centers. Generalization to contemporary critical care practice or non-U.S. settings is unknown. Validation on a contemporary cohort (e.g., MIMIC-III/IV) is the natural follow-up.

Fourth, our binary GBM at 60 days does not fully match SUPPORT-1995. Possible explanations include the model overlap (above), hand-crafted interaction terms in the original logistic regression, or insufficient hyperparameter tuning of our binary GBM. We have not pursued aggressive tuning because the central finding — that ranking-optimized survival ML underperforms — is robust to whether the matched binary classifier exactly closes the remaining gap.

Fifth, the demographic-fairness analysis (Section 3.9) is constrained by the size of the smallest strata (other race, n = 73; Black, n = 217), so those subgroup estimates are correspondingly uncertain; and, like all results here, it derives from a single historical U.S. cohort.

Sixth, the conformal procedure we apply uses simple right-censoring exclusion at the calibration step (Section 2.7); a more sophisticated alternative is the censoring-aware procedure of Candès et al. [25] with inverse-censoring-probability weighting. For SUPPORT2 at t = 60 and 180 days, no censoring exists before either horizon, so the two methods coincide.

Seventh, although the model panel is broad — Cox PH and elastic net, random survival forest, GBM Survival, DeepSurv, the discrete-time models nnet-survival and DeepHit, a horizon-matched binary GBM, and an MLP — every model is trained and tested on a single historical U.S. cohort (1989–1994). External validation on a contemporary cohort (MIMIC-IV, eICU) is the principal remaining step; we provide a ready-to-run adapter (`scripts/external_validation.py`) that applies the SUPPORT2-trained models, with identical preprocessing, to any cohort mapped to the SUPPORT2 schema, but credentialed external data were outside the scope of this reanalysis.

### 4.5 Conclusion

When ranking-optimized survival models are evaluated at the bedside — that is, against fixed-time mortality prediction rather than time-ranking — they can be outperformed by physicians and by decades-old logistic regression. This is not because the models are poorly calibrated; it is because their training objective is misaligned with the deployment target. Fixing the misalignment via timepoint-matched binary classification partially closes the gap. Leave-one-disease-out validation reveals severe generalization failures in the most distinct patient subgroups that are masked by the overall C-index. Mondrian-conformal intervals support principled selective deployment, with informative-tier patients achieving AUC ≥ 0.84 at both 60- and 180-day horizons. Sensitivity to missing-physician imputation reveals that the apparent physician advantage is partly selection-driven. Future clinical survival ML should report timepoint-specific metrics, train for the deployment target, validate across subgroups, quantify uncertainty rigorously, and account explicitly for missingness patterns in human comparators.

## Data and code availability

The SUPPORT2 dataset is publicly available at https://hbiostat.org/data. The complete reanalysis pipeline (nine checkpointed, resumable stages), all figures and tables, the full random-seed configuration, and every auxiliary analysis are openly available at https://github.com/ClevixLab/Ranking-optimized-survival-models under an MIT license (a Zenodo archive with a citable DOI accompanies the release). Anyone can clone the repository and reproduce every figure, table, and number: reproducibility is enforced by an automated verifier — make reproduce regenerates all core outputs from the raw data under a fixed seed, and make verify asserts each seed-deterministic headline result against a committed source-of-truth (tests/expected_results.json), exiting non-zero on any drift; the same check runs in continuous integration on every commit. A step-by-step map from each result in this paper to the exact command that produces it is given in Appendix A (Table A1).

## Ethics statement

This study is a secondary analysis of the SUPPORT2 dataset, which is publicly available in fully de-identified form (https://hbiostat.org/data). No new data were collected and no identifiable patient information was accessed; the work therefore did not require additional institutional review board approval or informed consent. The original SUPPORT study was approved by the institutional review boards of the participating centers [1].

## Author contributions

This study used the CRediT taxonomy. **Conceptualization:** Truong Quynh Hoa. **Methodology:** Truong Quynh Hoa. **Software:** Truong Quynh Hoa, Luu Duc Trung. **Validation:** Hoang Dinh Cuong, Truong Quynh Hoa. **Formal analysis:** Truong Quynh Hoa. **Investigation:** Truong Quynh Hoa, Hoang Dinh Cuong, Luu Duc Trung. **Data curation:** Luu Duc Trung. **Writing – original draft:** Truong Quynh Hoa. **Writing – review & editing:** Truong Quynh Hoa, Hoang Dinh Cuong, Luu Duc Trung. **Visualization:** Truong Quynh Hoa. **Supervision:** Truong Quynh Hoa. **Project administration:** Truong Quynh Hoa. Hoang Dinh Cuong led clinical and medical-content validation and clinical interpretation, including review of the disease-subgroup findings; Luu Duc Trung supported data cleaning and model execution with Hoang Dinh Cuong, under Truong Quynh Hoa’s direction including the selection of statistical tests. All authors reviewed and approved the final manuscript.

## Competing interests

The authors declare no competing interests.

## Funding

This work received no external funding.

## Data Availability

No new data were generated in this study. The SUPPORT2 dataset used in this study is publicly available from the GitHub repository of Gensheimer and Narasimhan at https://github.com/MGensheimer/nnet-survival/tree/master/data. All analysis code is openly available at https://github.com/ClevixLab/Ranking-optimized-survival-models

## Acknowledgments

We thank the original SUPPORT investigators for releasing the cohort publicly, and the maintainers of the scikit-survival library for the time-dependent AUC implementation. We acknowledge use of Claude (Anthropic) for code review and manuscript preparation; all analytical decisions and numerical results were verified by the authors.

## Appendix A: Reproducibility

All results in this paper are reproducible from the public repository (https://github.com/ClevixLab/Ranking-optimized-survival-models). After cloning, the core pipeline is regenerated and machine-verified with four commands:

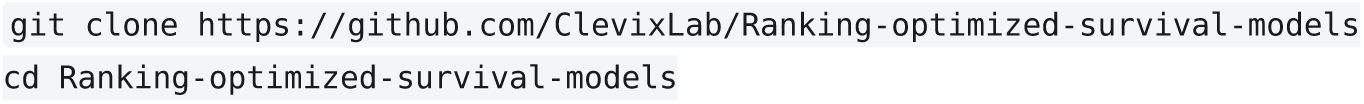
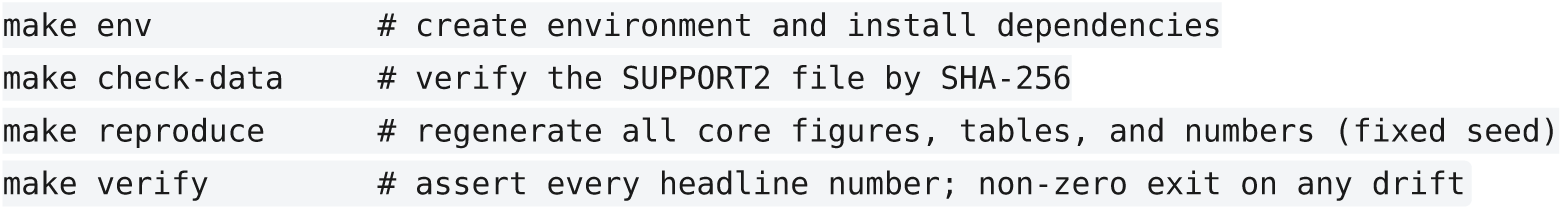

The optional deep-survival and auxiliary analyses (which require PyTorch) are run as standalone scripts:

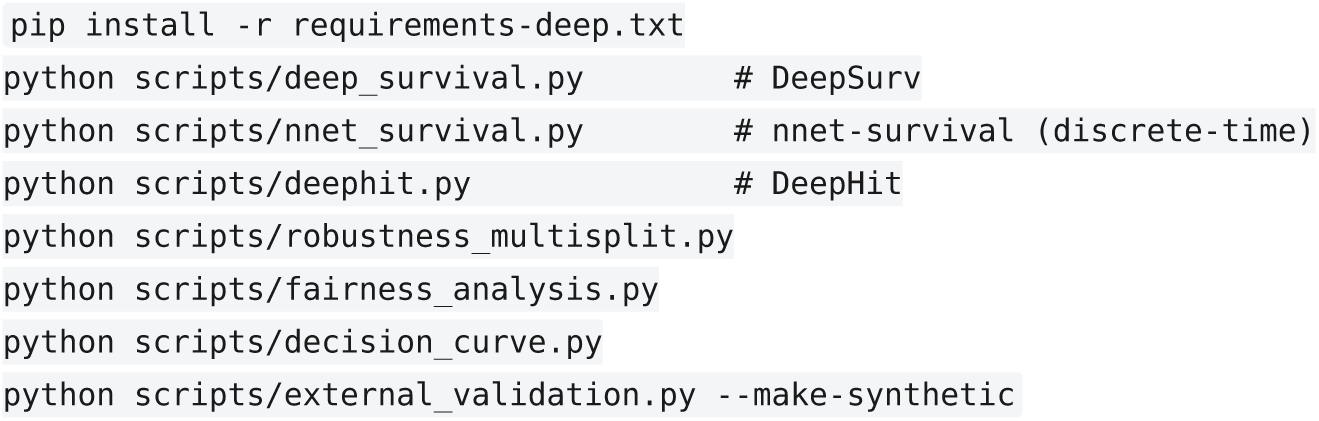

**Table A1.** Map from each reported result to the script/command that produces it. Core pipeline results are seed-deterministic and asserted by make verify; deep-learning models are marked platform-sensitive and are reported for completeness rather than bit-exact reproduction.

| Result in the paper | Produced by | Reproducibility |
| --- | --- | --- |
| Cohort & baseline C-indices (3.1); fixed-horizon AUCs (3.2); recalibration (3.3); cross-horizon matrix (Fig 5); five-predictor summary (Fig 6); binary/MLP fix (3.4); Figs 1-8; Tables 2-3 inputs | `make reproduce` → `scripts/pipeline_orchestrator.py` (stages 1-8) | seed-deterministic; `make verify` |
| Headline-number contract | `make verify` → `scripts/verify_reproduction.py` vs `tests/expected_results.json` | exits non-zero on drift |
| GBM hyperparameter sensitivity sweep (3.1) | pipeline (module 2b sweep) | seed-deterministic |
| Leave-one-disease-out BSS + CIs (3.5, Fig 7) | pipeline module 3 | seed-deterministic |
| Conformal coverage & informative-tier AUC (3.6, Fig 8) | pipeline module 4 | seed-deterministic |
| Missing-physician sensitivity (3.7, Table 2) | pipeline module 5 → `tables/table5_sensitivity.csv` | seed-deterministic |
| Multi-split robustness (3.8, Fig 9) | `python scripts/robustness_multisplit.py` | seed-deterministic |
| Demographic fairness (3.9, Table 3, Fig 10) | `python scripts/fairness_analysis.py` | seed-deterministic |
| Decision curve analysis (3.10, Fig 11) | `python scripts/decision_curve.py` | seed-deterministic |
| DeepSurv (3.1, 3.4) | `python scripts/deep_survival.py` | platform-sensitive (PyTorch) |
| nnet-survival (3.4 panel) | `python scripts/nnet_survival.py` | platform-sensitive (PyTorch) |
| DeepHit (3.4 panel) | `python scripts/deephit.py` | platform-sensitive (PyTorch) |
| External validation (4.4) | `python scripts/external_validation.py` (`--make-synthetic` or `--external <csv>`); see `docs/external_validation_MIMIC.md` | harness; external data user-supplied |

The exact software versions are pinned in `requirements.txt` (and `requirements-deep.txt` for the PyTorch models); the dataset is validated by SHA-256 before any computation.

Full reproducibility package and automated verifier: see the repository `README.md` and `REPRODUCIBILITY.md`. Reproduce end-to-end with `make reproduce && make verify`.

## Notes

### Competing Interest Statement

The authors have declared no competing interest.

### Funding Statement

The author(s) received no specific funding for this work.

### Author Declarations

This study used de-identified, publicly available data from the SUPPORT2 (Study to Understand Prognoses, Preferences, Outcomes, and Risks of Treatment) dataset, originally collected under informed consent at five US medical centers (1989–1994) and subsequently released for open research use. No new patient data were collected and no interaction with human participants was conducted as part of this study. Institutional review board (IRB) approval was therefore not required.

